# The potential health and economic value of SARS-CoV-2 vaccination alongside physical distancing in the UK: transmission model-based future scenario analysis and economic evaluation

**DOI:** 10.1101/2020.09.24.20200857

**Authors:** Frank Sandmann, Nicholas G. Davies, Centre for the Mathematical Modelling of Infectious Diseases COVID-19 working group, Anna Vassall, W John Edmunds, Mark Jit

**Affiliations:** London School of Hygiene and Tropical Medicine, London, UK; Statistics, Modelling and Economics Department, National Infection Service, Public Health England, London, UK; School of Public Health, University of Hong Kong, Hong Kong SAR, China

**Keywords:** SARS-CoV-2, COVID-19, vaccination, mathematical model, economic evaluation

## Abstract

**Background:** In response to the coronavirus disease 2019 (COVID-19), the UK adopted mandatory physical distancing measures in March 2020. Vaccines against the newly emerged severe acute respiratory syndrome coronavirus 2 (SARS-CoV-2) may become available as early as late 2020. We explored the health and economic value of introducing SARS-CoV-2 immunisation alongside physical distancing scenarios in the UK.

**Methods:** We used an age-structured dynamic-transmission and economic model to explore different scenarios of immunisation programmes over ten years. Assuming vaccines are effective in 5-64 year olds, we compared vaccinating 90% of individuals in this age group to no vaccination. We assumed either vaccine effectiveness of 25% and 1-year protection and 90% re-vaccinated annually, or 75% vaccine effectiveness and 10-year protection and 10% re-vaccinated annually. Natural immunity was assumed to last 45 weeks in the base case. We also explored the additional impact of physical distancing. We considered benefits from disease prevented in terms of quality-adjusted life-years (QALYs), and costs to the healthcare payer versus the national economy. We discounted at 3.5% annually and monetised health impact at £20,000 per QALY to obtain the net monetary value, which we explored in sensitivity analyses.

**Findings:** Without vaccination and physical distancing, we estimated 147.9 million COVID-19 cases (95% uncertainty interval: 48.5 million, 198.7 million) and 2.8 million (770,000, 4.2 million) deaths in the UK over ten years. Vaccination with 75% vaccine effectiveness and 10-year protection may stop community transmission entirely for several years, whereas SARS-CoV-2 becomes endemic without highly effective vaccines. Introducing vaccination compared to no vaccination leads to economic gains (positive net monetary value) of £0.37 billion to +£1.33 billion across all physical distancing and vaccine effectiveness scenarios from the healthcare perspective, but net monetary values of physical distancing scenarios may be negative from societal perspective if the daily national economy losses are persistent and large.

**Interpretation:** Our model findings highlight the substantial health and economic value of introducing SARS-CoV-2 vaccination. Given uncertainty around both characteristics of the eventually licensed vaccines and long-term COVID-19 epidemiology, our study provides early insights about possible future scenarios in a post-vaccination era from an economic and epidemiological perspective.

**Research in Context:** *Evidence before this study:* We searched PubMed and medRxiv for economic evaluations of SARS-CoV-2 vaccines with the search string (coronavirus OR COVID OR SARS-CoV-2) AND (vaccin* OR immunisation) AND ((economic evaluation) OR (cost effectiveness analysis)) AND 2020[dp] on September 21, 2020, with no language restrictions. We found one pre-print that valued health outcomes in monetary terms and explored the additional impact of vaccines in a cost-benefit analysis of physical distancing for the USA; no study focused on vaccines in a full economic evaluation.

*Added value of this study:* With a growing number of vaccine candidates under development and having entered clinical trials, our study is to our knowledge the first to explore the health and economic value of introducing a national SARS-CoV-2 immunisation programme. A programme with high vaccine effectiveness and long-lasting protection may stop the community transmission entirely for a couple of years, but even a vaccine with 25% vaccine effectiveness is worthwhile to use; even at short-lived natural and vaccine-induced protections. After an initial lockdown, voluntary physical distancing as a sole strategy risks a large second epidemic peak, unless accompanied by highly effective immunisation. Compared to no vaccination, introducing vaccination leads to positive net monetary value across physical distancing scenarios from the healthcare perspective, subject to the long-run vaccine price and cost-effectiveness of other treatments (e.g. new drugs). The net monetary value of immunisation decreases if vaccine introduction is delayed, natural immunity is long or vaccine-induced protection is short. Intermittent physical distancing leads to negative net benefits from the perspective of the wider economy if the daily national income losses are persistent and large.

*Implications of all the available evidence:* Our model findings highlight the health and economic value of introducing SARS-CoV-2 vaccination to control the COVID-19 epidemic. Despite the many uncertainties, continued physical distancing may be needed to reduce community transmission until vaccines with sufficiently high vaccine effectiveness and long-lasting protection are available. Our study provides first broad health-economic insights rather than precise quantitative projections given the many uncertainties and unknown characteristics of the vaccine candidates and aspects of the long-term COVID-19 epidemiology, and the value of vaccines will ultimately depend on other socioeconomic and health-related policies and population behaviours.

## Introduction

In early 2020, physical distancing (or “social distancing”) measures were adopted in at least 149 countries in response to the widespread community transmission of the newly emerged severe acute respiratory syndrome coronavirus 2 (SARS-CoV-2).^1,2^ Physical distancing measures included school, non-essential retail, hospitality venues and workplace closures; orders to stay at home and keeping a safe distance of 1-2 meters from individuals from different households; restrictions on mass gatherings and movement (including public transport); and isolation of symptomatic cases and quarantining exposed individuals.^2^ The UK national government adopted similar measures in March 2020 following a rapid rise in cases of coronavirus disease 2019 (COVID-19) and the prospect of the healthcare system becoming overwhelmed.^3^

The mandatory measures of physical distancing were implemented in the early stages of the pandemic on the basis of the high transmissibility of SARS-CoV-2 and burden of COVID-19, uncertainties about the COVID-19 epidemiology,^1^ the absence of effective pharmaceutical interventions against COVID-19, and the imperative to save lives under the rule of rescue.^4^ Nonetheless, the pandemic response has led to large contractions of the global economy, and the largest contraction in the UK economy since monthly records began in 1997, with gross domestic product (GDP) falling by 5.8% in March 2020 and by 20.4% in April 2020.^5^ So far, COVID-19 and associated policy responses have cost the UK’s economy at least £192 billion in 2020-2021, which is nearly 10% of the annual gross domestic product,^6^ without including the full macro-economic impact of both response and disease.

In terms of minimising health losses and economic harm, apart from continued non-pharmaceutical interventions like physical distancing and extended testing and tracing for SARS-CoV-2 infection, effective and safe pharmaceutical interventions will be required. As of 21 September 2020, the World Health Organization is tracking 149 vaccine candidates in preclinical evaluation and 38 candidate vaccines are in clinical trials of which nine candidate vaccines are in phase-3 trials.^7^ Some countries have signed advanced purchasing agreements for early access to vaccines at initial emergency cost-prices and shared financial risks.^8^ The UK has signed agreements for at least six vaccine candidates and co-funds clinical trials, and the first vaccine may be available as early as in late 2020.^9^ Vaccination provides a potential control strategy not requiring recurring lockdowns or extensive testing, but their efficacy is unknown and hard to predict. Therefore, we aimed to explore the health and economic value of introducing a SARS-CoV-2 immunisation programme in the UK. Given the unknown characteristics of the eventually licensed SARS-CoV-2 vaccines, and the longer-term epidemiology of COVID-19, we focussed on the wider impact of vaccination alongside physical distancing scenarios as the value of vaccines will depend on other policies and population behaviours (both economic and health related).

## Methods

### Epidemiological model

We used CovidM, an age-stratified dynamic transmission model that was developed to explore the effect of COVID-19 and non-pharmaceutical interventions in the UK.^10^ In this SEIR (susceptible, exposed, infectious and recovered) epidemic model, individuals are stratified into 5-year age bands and susceptible individuals may become infected and move into an exposed compartment before becoming infectious and symptomatic. A proportion of individuals will also be asymptomatically infected only. Individuals recover but natural protection may wane over time. Our estimated cases refer to symptomatic (clinical) cases. The model also tracks COVID-19-related hospitalisations and deaths by age. For more details, see Davies et al. (2020).^10^ We also explored updating parameters using data from the UK (appendix, pp. 2-3).

We extended the compartmental model to investigate the health impact of an immunisation programme and the economic value of SARS-CoV-2 vaccination in the UK. Individuals who were vaccinated move into separate compartments, assuming all-or-nothing protection from infection, and they may also lose the vaccine-induced protection exponentially over time. We added demography in terms of births and (disease-unrelated) deaths, which allows exploring outcomes over ten years (2020-2029) to sufficiently capture the potential effects of vaccination.

The model starts with a basic reproductive number, R_0_, of 2.7 (95% credible interval: 1.6, 3.9).^10^ At the start of the model and every month afterwards, five new infections are introduced into the population each day for seven days in the age groups 20-49 years to represent a low but continuous importation of infections.^11^

### Immunisation programme intervention and comparator scenarios

The model compared (A) no-vaccination baseline scenario with two vaccination programmes resembling pessimistic and optimistic extremes of a reasonable spectrum of assumptions about vaccine characteristics; (B) Vaccination with 25% vaccine effectiveness (VE), vaccine-induced protection of 1-year duration, and annual revaccination of 90%; and (C) Vaccination with 75% VE, vaccine-induced protection of 10-year duration, and annual revaccination of 10% of individuals in the target population. Initially, in both vaccination scenarios we assumed a vaccination rate of 1 million individuals a day until uptake reaches 90%, which was informed by the stock of doses procured in early contracts of the UK government and assuming vaccination to be prioritised in healthcare facilities (potentially with the help of other qualified personnel like pharmacists). Vaccination was assumed to prevent transmission and disease. Vaccination was targeted with uniform coverage at individuals aged 5-64 years, which is largely overlapping with the age range of participants in clinical trials of vaccine candidates and also including school children.^7^ If individuals aged >64 years can be vaccinated they will become a potential target group in future because of their high risk of severe disease. Natural protection from infection was assumed to last for a mean of 45 weeks in the base case.^12^ In sensitivity analyses we varied the duration of natural and vaccine-induced protection from 12 weeks to 10 years.

We assumed vaccination introduction starting from 01 December 2020.^13^ In a separate sensitivity analysis, we explored the change in the net monetary value of vaccination if vaccine introduction is delayed up until the start of 2022.

### Physical distancing scenarios

The model considered the vaccination scenarios with and without physical distancing. Physical distancing was implemented by reducing the number of close-contacts of individuals at home, work, school, and other locations (such as transport and leisure; appendix, p. 1). We consistently assumed voluntary physical distancing of the public after the initial outbreak by keeping a safe space from individuals from other households, face coverings, increased hand hygiene, and working from home, with close-contacts being reduced by 10% at work, school, and other locations. All scenarios also considered summer and winter holidays, during which school contacts were reduced to 0 (appendix, p. 1). The model made no explicit assumptions about increased mixing between children from different schools and locations during holidays e.g. during play dates.

We then explored three groups of physical distancing scenarios of either (i) no historical lockdown (a counterfactual scenario); (ii) an initial lockdown only as observed historically; and (iii) intermittently occurring physical distancing where the contact rates are reduced once incidence goes over a certain level (varied in sensitivity analysis to be triggered at incidence levels of 10 to 60 cases per 100,000 population). Technically, the initial lockdown and the intermittently occurring physical distancing result in a similar scaling of the contact matrices (appendix, p. 1). Contacts are increased again once the daily incidence reaches <500 cases (about 1/100,000 cases). All simulations started on 01 January 2020 for 10 years.

### Comparison of estimated with observed hospitalisations

For scenarios with an initial lockdown, the incidence threshold for the initial lockdown and physical distancing was based on aligning the estimated cumulative number of hospitalisations to the observed numbers in the UK in mid-July 2020 (appendix, p. 1), with an estimated 8.54% infections by mid-July 2020. Rather than to reproduce the first few months of the epidemic precisely, however, we did not fit the model to the full range of data available or explored targeted vaccination strategies given the many uncertainties and unknown characteristics of the vaccine candidates and aspects of the COVID-19 epidemiology.

### Health and economic impact

The economic analysis was fully integrated with the epidemiological model. Our main analysis adopted the reference case used to evaluate vaccines in the UK,^14,15^ considering benefits in terms of disease prevented using quality-adjusted life-years (QALYs), and the costs from the perspective of the National Health Service (NHS) over a ten-year timeframe. The main outcome was the net monetary value of a scenario, i.e. the combined health (i.e. QALY) and economic benefits of the interventions expressed in monetary terms (QALY * monetary_value_per_QALY - cost).^16^ We assumed a monetary value per QALY of £20,000,^14,15^ which we varied between £0 and £60,000 in sensitivity analysis.^17^ Note that positive incremental net monetary values of vaccination against no-vaccination thus indicate results within the cost-effectiveness threshold of e.g. £20,000/QALY (i.e. vaccination and/or physical distancing is cost-effective), while negative incremental net monetary values indicate results above the threshold.^16^ Future costs and QALYs were discounted at 3.5% annually,^14,15^ and we explored varying the discount rate between 0%-10% in a sensitivity analysis.^14,17^

For the health impact, we considered the QALYs lost by symptomatic cases, non-fatal hospitalisations, intensive-care unit (ICU) survivors, adverse-events following immunisation (AEFI), and premature fatalities due to COVID-19 (appendix, p. 4). For the costs from the NHS perspective, we considered the expenditures on non-fatal hospitalisations (ICU and non-ICU), enhanced personal protective equipment, visits to general practitioners, remote helpline calls, AEFI, vaccine administrations, and the vaccine costs (appendix, pp. 5-6). We assumed a conservative long-run cost-price per dose of £10 (similar to influenza vaccines), which we varied in sensitivity analyses at £0-£200 (with £0 representing the value of the vaccine alone). Additional costs of setting up a delivery programme have not been included. In addition, the vaccination scenarios considered the public expenditures on subsidising the development of SARS-CoV-2 vaccines with £250 million by the UK government so far,^13^ which could be regarded as an extraordinary lump-sum ex-ante premium.

We conducted an exploratory, secondary analysis that considered the negative impact of physical distancing for the wider national economy, and the incremental benefit of introducing vaccination on that impact. This analysis may be used as a framework until we know more about the vaccines. We approximated the costs of physical distancing in terms of losses to the UK’s GDP, which is a monetary output measure of all goods and services produced in a country during a specific period. We assumed that daily GDP was £5,757 million based on the seasonally-adjusted GDP in 2019, Q4, of £523,917 million,^18^ and we assumed national income losses from physical distancing and potentially other measures aiming to reduce transmission. GDP losses were assumed for each day the incidence exceeded 1,000 new cases due to the 10% reduced contacts from voluntary physical distancing, with the equivalent of 2% of the daily GDP (similar to the largest pre-COVID-19 reduction since 1997; the 1,000 new cases were approximated by the numbers in mid-March 2020).^5^ Losses on days of the scenarios with an initial lockdown and subsequent physical distancing were evaluated against the same threshold. However, for days with intermittently occurring physical distancing at incidence levels of 10 to 60 cases per 100,000 population, we additionally explored higher GDP losses between 0% and 20% (similar to the monthly fall in GDP of 5.8% in March 2020 and 20.4% in April 2020).^5^ We did not add health sector costs estimated under the NHS perspective or productivity losses in COVID-19 cases to these wider economic costs, to avoid double counting.

To account for parameter uncertainty, we ran the epidemiological model deterministically with R0 values of 2.7 (the base case) as well as 1.6 and 3.9.^10^ The economic model used a probabilistic sensitivity analysis with 1,000 iterations (appendix, pp. 3-5).

## Results

### Health outcomes and disease dynamics

An unmitigated epidemic without a lockdown is expected to have led to high incidence levels in the initial outbreak (Figure 1a), with smaller but recurring annual outbreaks due to varying contact rates and transmission during holidays, loss of natural immunity and births replenishing the susceptible population. In contrast, implementing an initial lockdown only as observed historically moves the high burden into the second peak (Figure 1b), with little change in the overall burden as compared to no lockdown over 10 years (Figure 2a). Intermittent physical distancing in future may change the height of future peaks and splits up the epidemic into two smaller outbreaks each year, with a consequent reduction in burden (Figure 1c-f).

**Figure 1:**
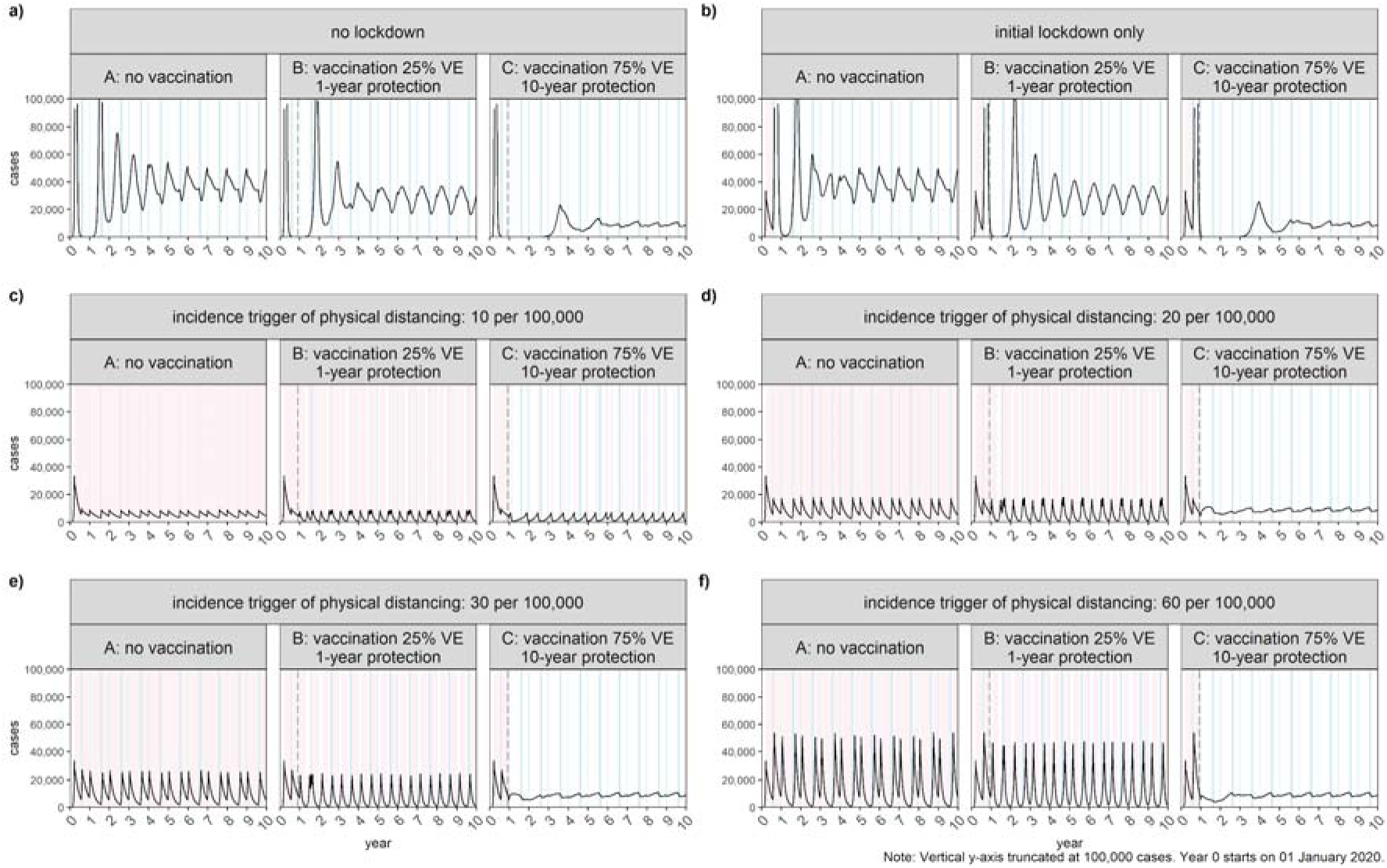
Epidemiological impact of vaccination. Time-series of the daily number of symptomatic cases for selected physical distancing scenarios (physical distancing trigger at incidences of 40 and 50 per 100,000 not shown for ease of presentation). Days highlighted indicate summer and winter holidays (in blue), periods of physical distancing (red), or neither of the two (white). Note: The y-axis is truncated at 100,000 cases daily to allow meaningful visual comparisons across panels.

**Figure 2:**
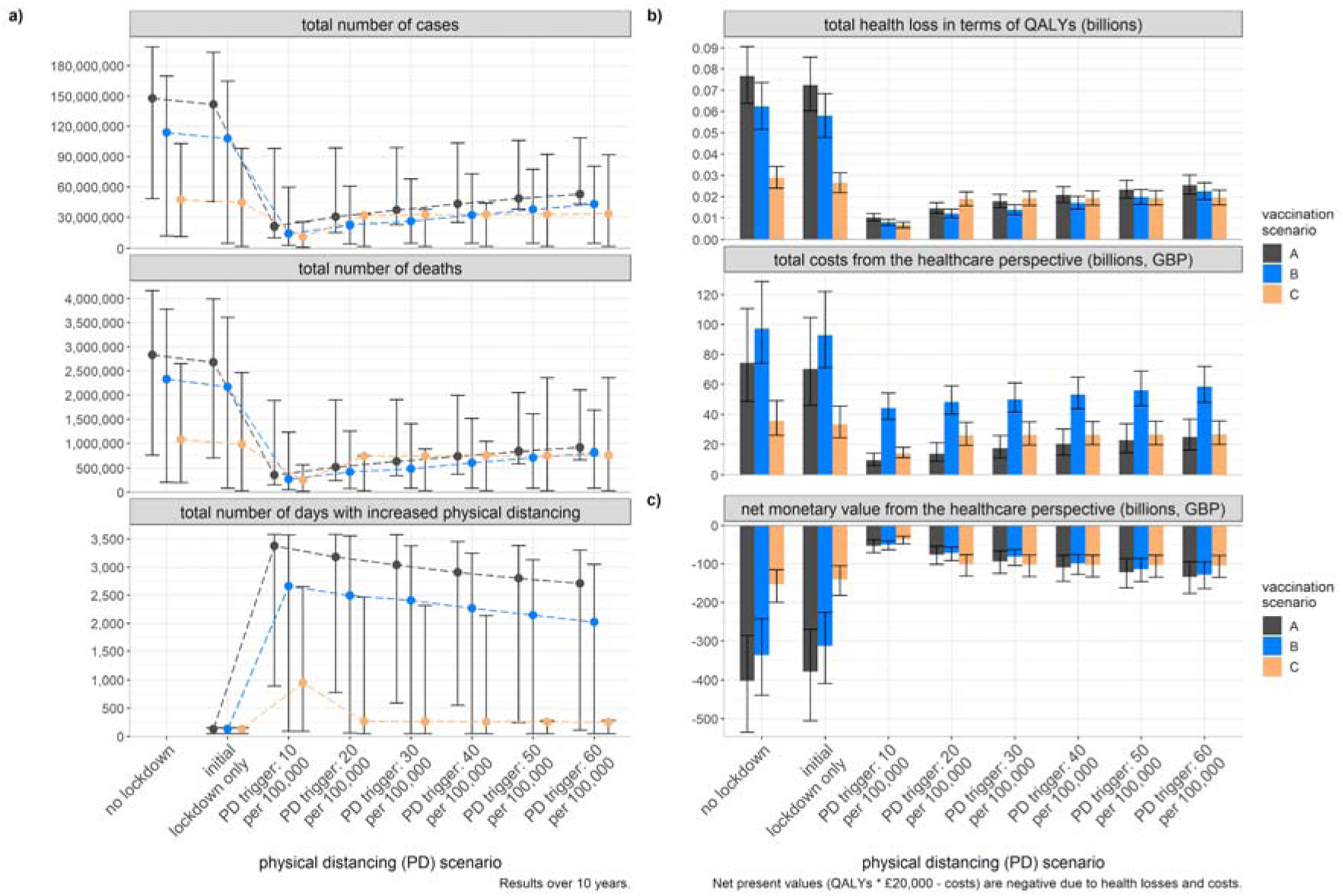
Health and economic impact of vaccination. Results over ten years for different physical distancing scenarios. Panel a) shows the estimated numbers of cases, deaths, and days of physical distancing; panel b) shows the total costs and QALYs lost; panel c) shows the net monetary value (healthcare perspective).

Introducing vaccination may lower and delay the peaks of future outbreaks (Figure 1). Highly effective vaccines may reduce transmission to a bare minimum for 1-2 years, while SARS-CoV-2 is likely to become endemic (Figure 1).

The 10-year burden of an unmitigated epidemic in the UK (the counterfactual scenario) was estimated with 147.9 million cases (95% uncertainty interval: 48.5 million, 198.7 million), 2.8 million deaths (767,000, 4.2 million), and 76.7 million QALYs lost (63.8 million, 90.7 million). Combined with intermittent physical distancing, these numbers can be reduced over the same time period to 21.4-53.0 million cases, 0.36-0.92 million deaths, and 10.1-25.4 million QALYs lost (appendix, pp. 7-8). Introducing vaccination is estimated to reduce these numbers further to 11.5-43.1 million cases, 0.25-0.82 million deaths, and 6.7-22.5 million QALYs lost (Figure 2a-b). The estimated number of days in lockdown or with increased physical distancing is substantial, unless vaccines with high VE and long-lasting protection become available (Figure 2a-b).

### Costs and net monetary value for healthcare perspective and wider national economy

The total costs from the healthcare perspective over ten years are highest in the unmitigated scenario without vaccination and physical distancing, resulting in an absolute net monetary value of -£402 billion (−286 billion, −535 billion). Compared to no vaccination, introducing vaccination always leads to positive incremental net monetary value across physical distancing scenarios between £0.37 billion and £1.33 billion (Figure 2b).

The incremental net monetary value of introducing vaccination versus no vaccination is also always positive across physical distancing scenarios from a wider economic perspective (Figure 3a-d). With a lower incidence rate threshold for physical distancing, however, greater losses of national income are expected that lead to negative net monetary value from the perspective of the wider national economy if the (proportion of) daily GDP losses are high (Figure 3); note that the equivalent loss may be different per physical distancing scenario (e.g. losses may be high and closer to 15% at a lower incidence threshold of 10/100,000 cases).

**Figure 3:**
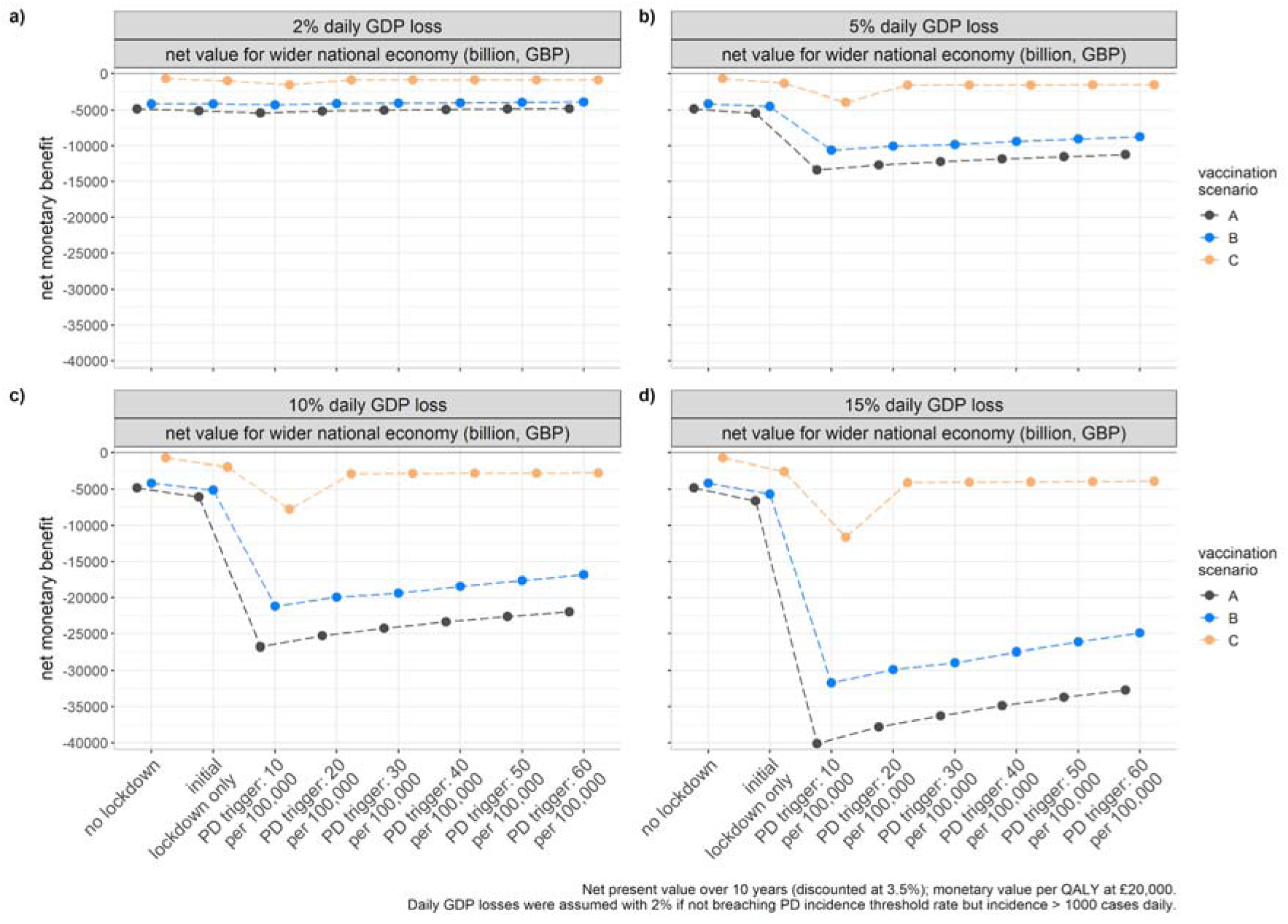
Wider economic impact of vaccination. The potential wider economic impact of vaccination in terms of net monetary values for different (proportions of) daily GDP loss during physical distancing (PD).

### Sensitivity analyses

The economic value of introducing vaccination was estimated to be lower if the vaccine is introduced in December 2020 compared to an earlier introduction (Figure 4). The incremental net monetary value of introducing vaccination does not always decrease as the delay increases, due to the interaction with seasonal cycles in incidence, holidays, and periods of physical distancing.

**Figure 4:**
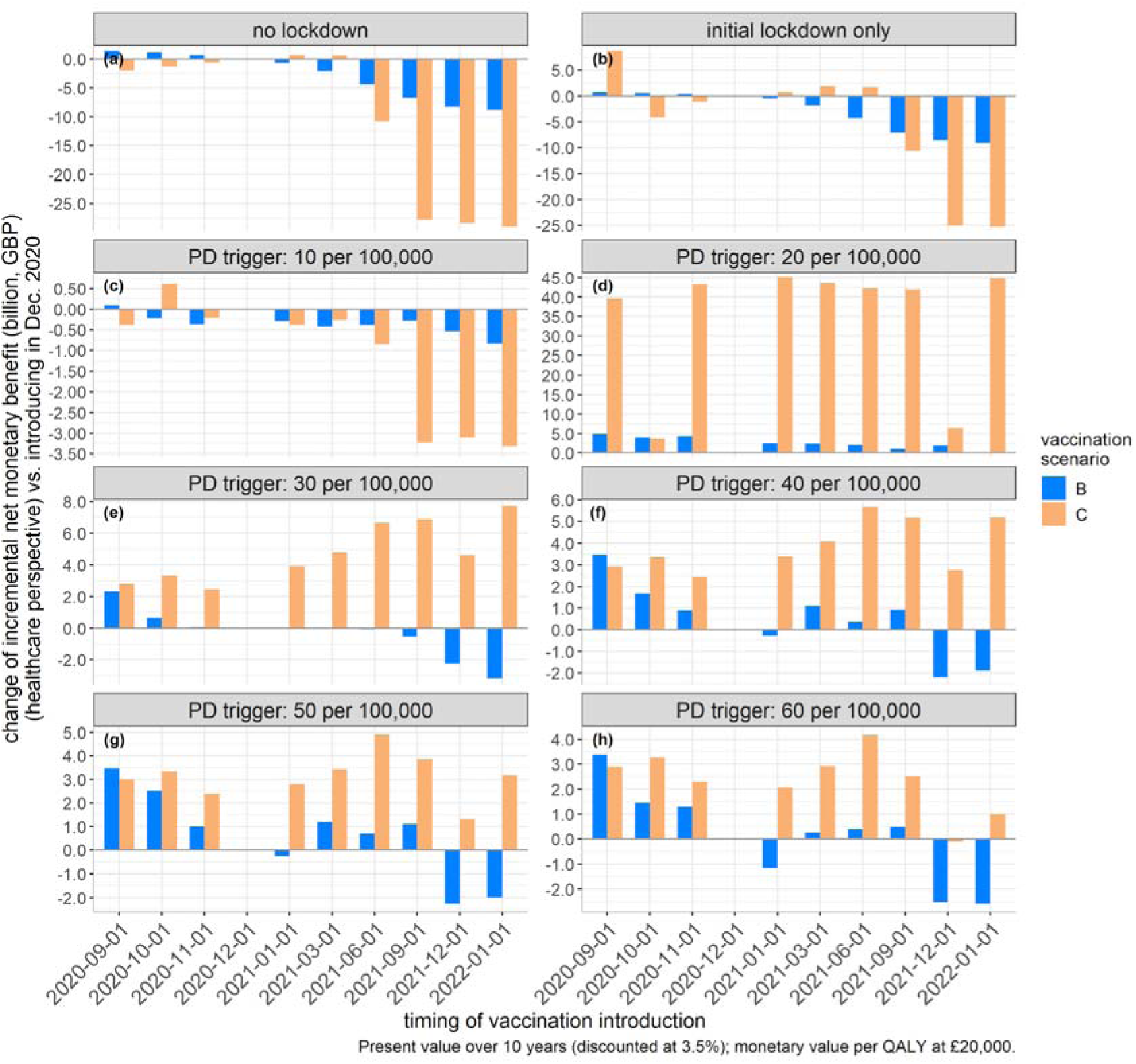
Timing of vaccination. Sensitivity analysis on the timing of vaccination introduction in terms of changes to the net monetary benefit when introducing vaccination in December 2020.

With a longer duration of natural immunity, the economic value of immunisation decreases (Figure 5a), and it may indeed become lower than no vaccination for long durations of natural immunity (5-10 years; Figure 5a, c-e). Similarly, with a longer duration of vaccine-induced protection the economic value of vaccination compared to no vaccination increases, but the impact depends on the vaccine characteristics of the two vaccination scenarios regarding e.g. VE and coverage (Figure 5b).

**Figure 5:**
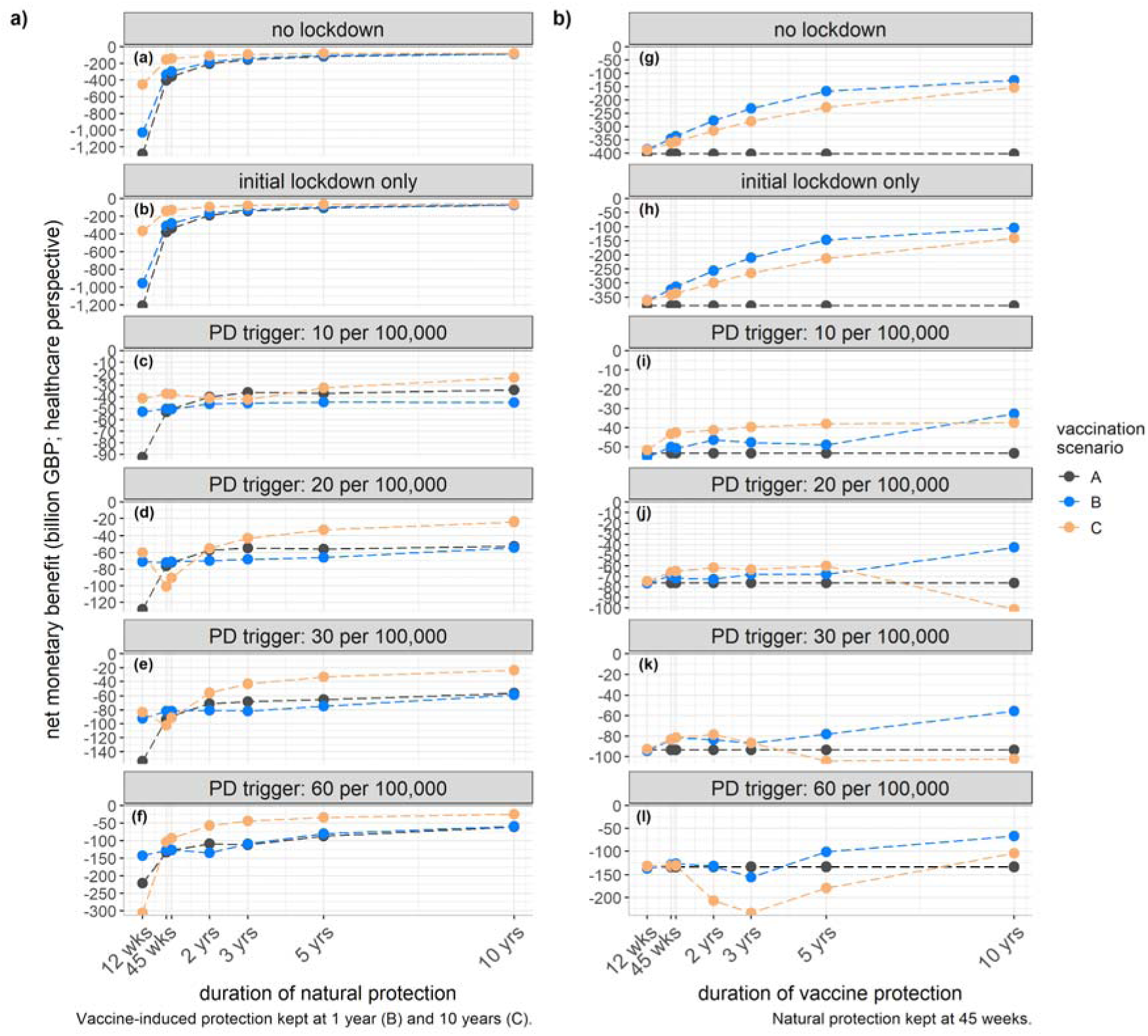
Natural and vaccine-induced immunity. Sensitivity analysis on the duration of natural protection (panel a) and vaccine-induced protection (panel b) against SARS-CoV-2 infection

The base-case findings were also largely robust when using different values for the monetary value of a QALY (Figure 6a) or discount rates (Supplementary Figure 1). From the healthcare perspective, a viable price per dose in future will depend on the vaccine characteristics and other policies and population behaviours (Figure 6b).

**Figure 6:**
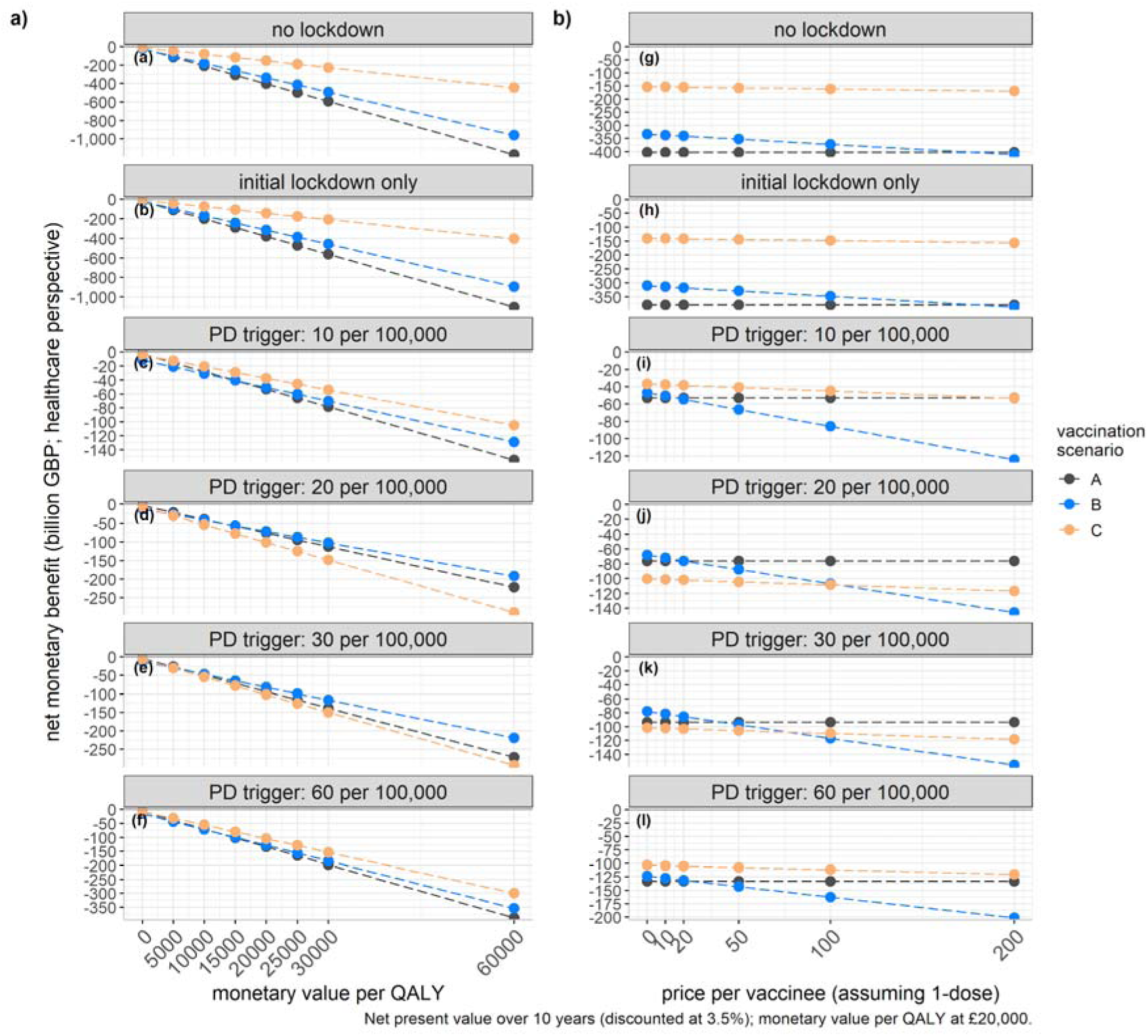
Value of quality-adjusted life year. Sensitivity analysis on the monetary value per QALY (panel a) and the price per vaccine (panel b).

**Supplementary Figure 1:**
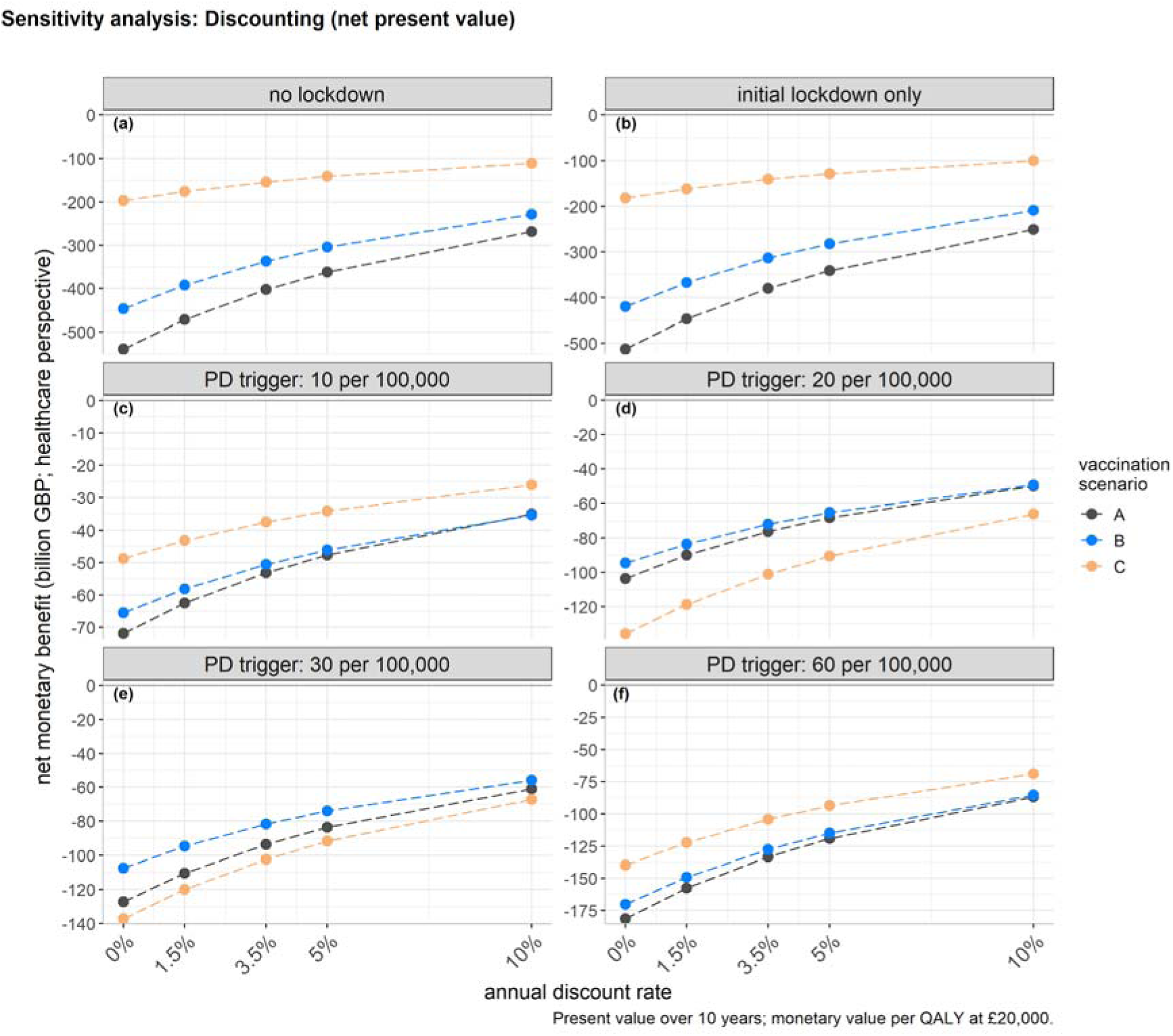
Sensitivity analysis on the discount rate.

## Discussion

This study explored the health and economic impact of introducing a SARS-CoV-2 vaccination programme in the UK. Our model findings show that highly effective vaccines with long-lasting protection can reduce transmission over the next 10 years, while SARS-CoV-2 is likely to become endemic. Compared to no-vaccination, introducing vaccination leads to positive net monetary value across physical distancing scenarios. Even in a pessimistic scenario in which SARS-CoV-2 vaccines have limited efficacy and provide only short-lived protection, their use would still offer overall benefit given the large health and economic costs of COVID-19. The value of the vaccine to the UK alone could be in the billions and possibly trillions of pounds. This indicates that investments in SARS-CoV-2 vaccination have been worthwhile to accelerate the development of vaccine candidates to make the vaccines available at accessible prices for both the UK as well as other countries, instead of competing at premium monopoly prices.

Vaccination introduction by the end of 2020 seems an ambitious goal; it demands an unprecedented speed of development for a licensed pharmaceutical intervention to be available within a year after the targeted disease was first described.^1,19^ The UK government has signed further contracts with other manufacturers to maximise the probability that at least one of the vaccine candidates will be successfully licensed by diversifying the risk.^9,19^ Within the framework of the analysis our model results indicate the continued importance of physical distancing to reduce community transmission in future, until a highly effective SARS-CoV-2 vaccination becomes available. Without such measures or other improvements e.g. in treatment, the initial lockdown will only have succeeded in delaying rather than mitigating a large epidemic. The value of subsequent physical distancing is even greater than reflected in the total QALYs and costs averted, since it splits annual epidemics into two separate peaks, which reduces the overall size of the peaks and hence the risk of exceeding hospital capacity. However, the intensity and economic cost of these subsequent periods of physical distancing may be less than those of the initial lockdown, if they can be combined with other measures such as effective testing and contact tracing, improvements of treatment, better availability of PPE, and precautionary behaviour such as wearing face coverings and handwashing by the general population. Furthermore, other considerations may become important such as the logistical challenge of administering a large quantity of doses within a short timeframe; possibly additional qualified healthcare workers could be recruited to administer the vaccines outside of primary care settings, but replacing the time that could have been used for alternative activities.

To our knowledge, this study is the first economic evaluation focussing on SARS-CoV-2 vaccination,^20^ and using established reference case methods.^14,15^ A few studies model the dynamics of SARS-CoV-2 vaccination (see e.g. ^21^). Although a conventional cost-effectiveness framework has its limitations during a pandemic, we find that even from the more conservative healthcare perspective the net benefits of vaccination are positive. Arguably, the monetary value per QALY could be different in this pandemic to the normal decision rules used in the UK given that (i) the value of the opportunity costs of COVID-19 admissions may be lower,^22^ (ii) we quantified the reduction in health loss and people generally prefer to avoid losses (loss aversion),^23^ and (iii) the health opportunity costs will change with COVID-19 given that the overall health sector budget may shift and COVID-19 will have an impact on the efficiency of other healthcare interventions. Some have also argued that a higher discount rate may be justified given that past policy decisions before the emergence of SARS-CoV-2 imply a relatively low value was placed on future non-influenza pandemics.^24^ We also highlight the potential magnitude of broader societal benefit, but given that our methods are simple, and the need for further macroeconomic modelling, we did not explore other future research avenues such as threshold pricing against a cost-effectiveness threshold or net societal benefit.

### Strengths and Limitations

We extended an age-structured dynamic-transmission model that was previously used to inform UK policies on introducing non-pharmaceutical interventions to provide early insights into the health and economic value of SARS-CoV-2 vaccination in the UK. Given the many uncertainties and unknown characteristics of the vaccine candidates and even the longer-term COVID-19 epidemiology, however, rather than to reproduce the first few months of the epidemic precisely we did not fit the model to the full range of data available. We also did not explore targeted vaccination strategies such as prioritising vaccination of key workers or high-risk individuals, and we did not include therapeutic pharmaceutical interventions that may not prevent transmission given our focus on vaccines (and which are unlikely to alter broad conclusions since most of the economic value of vaccination stems from averting non-hospitalised cases and damage to the wider economy). Thus, the main aim of our analysis was to generate qualitative insights, not precise numbers.^25^

In an exploratory secondary analysis our study also looked at the population-level trade-off in terms of health gains from disease averted versus costs to the economy. Although the modelled scenarios of physical distancing may not predict future disease dynamics for the next decade, physical distancing itself was not the focus of this study. However, ignoring the wider economic impact of physical distancing risks distorting conclusions as a lockdown may indeed help reduce the health burden and healthcare costs and thus look highly cost-effective from a healthcare perspective, but it will harm the wider economy and society from a broader perspective. We also did not account for (1) the economic impact of physical distancing causing long-term harm (such as reducing productivity) or conversely being mitigated by longer-term structural change (such as shifts in employment towards economic sectors more resilient to physical distancing measures), (2) that the actual GDP impact during the initial lockdown was mitigated in the UK by government actions that bear future cost implications, and (3) that governments may become more efficient at minimising health loss vs. economic trade-offs over time. Similarly, we did not account for future economic shocks. Although the analysis considered the impact for the UK population with 66.4 million individuals in England, Wales, Scotland, and Northern Ireland, the policy measures of physical distancing differ slightly between the four. Also, we did not address growing concerns about inequality, although an effective vaccine with high uptake will likely benefit subgroups that have suffered the worst health and economic losses from COVID-19.

Additional factors that were not considered in this analysis may become important in future, including enhanced testing programmes and other pharmaceuticals that may reduce the burden of disease and transmissibility of SARS-CoV-2. Some of these effects may have been included implicitly in our study by assuming long-term lower contact patterns of individuals following the first epidemic peak. Our economic analysis is also conservative in its input parameters, informing costs and quality-of-life values from other respiratory infections, which likely underestimates the QALY gain from preventing persistent COVID-19 symptoms.^26^ Moreover, our study ignored the health opportunity costs for displaced patients without COVID-19, excess mortality, and the longer-term impact on e.g. cancer care.^27^ Both the COVID-19 pandemic and the SARS-CoV-2 vaccination introduction may also disrupt healthcare service delivery of other vaccination programmes,^28,29^ and possibly the disease dynamics of other close-contact infections.^30^ We were also unable to provide accurate estimates on the wider social opportunity costs arising from the longer-term impact on mental health, household finances, and education both at an individual and societal level.^24^ Some of these difficulties stem from estimating how COVID-19 interacts with feedback loops in the health system as an external shock and will need to be quantified later.

In conclusion, our findings highlight the health and economic value of introducing SARS-CoV-2 vaccination to control the COVID-19 epidemic. Continued physical distancing may be needed to reduce community transmission until vaccines with high vaccine effectiveness and long-lasting protection are available. Our study provides broad insights rather than precise quantitative projections given the many uncertainties and unknown characteristics of the vaccine candidates and aspects of the long-term COVID-19 epidemiology, and the value of vaccines will ultimately depend on other policies and population behaviours (both economic and health related).

## Data Availability

Data were taken from published sources.

## Supplementary Material

### Physical distancing scenarios

For scenarios with an initial lockdown, the incidence threshold for the initial lockdown and physical distancing was based on aligning the results for the estimated numbers of COVID-19 related hospitalisations and deaths to the observed numbers in the UK. As of 15 July 2020, there were 130,472 recorded admissions and 41,035 deaths due to COVID-19 in the UK.^31^ The model most closely resembled these values when going into lockdown once the incidence reached 30 cases per 100,000 population and using the values shown in Supplementary Table 1, with an estimated 136,500 admissions and 34,055 deaths by 15 July 2020.

There are many unknowns surrounding the characteristics of the vaccine candidates, uncertain aspects of the longer-term COVID-19 epidemiology, what measures will be put in place in future, and the COVID-19-specific impact on costs and QALYs. However, in this economic evaluation a higher value was placed on approaching the absolute numbers as observed historically rather than closely resembling the observed disease dynamics, which are difficult to predict in future, too. Although the modelled scenarios of physical distancing may not perfectly predict future disease dynamics for the next decade, physical distancing itself was not the focus of this study evaluating the impact of SARS-CoV-2 vaccination. However, ignoring the wider economic impact of physical distancing risks distorting conclusions as an indefinite lockdown may indeed help reduce the health burden to a minimum and at minimal healthcare costs, but at substantial harm to the wider economy and society.

**Supplementary Table 1:** Reduced contacts between individuals who are physically distancing; these values were used to scale the underlying contact matrices obtained from POLYMOD.^32^

|  | home | work | school | other |
| --- | --- | --- | --- | --- |
| initially | 1.0 | 1.0 | 1.0 | 1.0 |
| physical distancing after the first (and subsequent) outbreak(s) (once daily incidence <500 cases, about <1/100,000 cases) | 1.0 | 0.67 | 0.67 | 0.67 |
| summer/winter holidays | 1.0 | 0.67 | 0.0 | 0.67 |
| initial lockdown (all scenarios except the “no lockdown” one) | 0.9 | 0.1 | 0.1 | 0.1 |
| subsequent physical distancing (triggered at incidence rates of 10-60/100,000 cases; released again once daily incidence <500 cases) | 1.0 | 0.1 | 0.1 | 0.1 |
| Note: The relative scaling of contacts during the initial lockdown was chosen to achieve a 75% reduction of $R_t$ as observed historically in the UK in 2020,<sup>33</sup> while closely matching observed admissions in the UK in mid-July 2020. Future summer and winter holidays were chosen in line with the dates of school holidays in England in 2020/2021. | | | | |

### Burden estimation

We considered the health burden of COVID-19 and related interventions in terms of symptomatic cases, non-fatal hospitalisations, intensive-care unit (ICU) survivors, adverse-events following immunisation (AEFI), and premature fatalities due to COVID-19. For the healthcare costs, we considered visits to general practitioners, remote helpline calls, non-fatal hospitalisations (ICU and non-ICU), enhanced personal protective equipment, AEFI, vaccine administrations, and the vaccine costs.

Key epidemiological illness parameters were taken from CovidM (Supplementary Table 2),^10^ which in turn informed these values by published estimates. For our study, we extended the model to incorporate demography in terms of births and (disease-unrelated) deaths to replenish susceptible individuals, assuming a death rate identical to the birth rate based on 731,000 live births in the UK in 2018,^34^ which allows exploring a longer timeframe over ten years (2020-2029). We also updated the proportion of inpatients who were admitted to critical care (0.17) and who died in critical care (0.32) based on a large study from the UK of more than 20,000 inpatients.^35^ Similarly, we used the age distribution of hospitalisations to inform the age-dependent proportion of hospital admissions in the UK.^35^

**Supplementary Table 2:** Input parameters for the epidemiological model

|  |  |  |  |
| --- | --- | --- | --- |
| Parameter | Value | Distribution | Sources |
| Latent period (E to I<sub>p</sub> and E to I<sub>s</sub>; days) | 4.0 | gamma | Davies et al. (2020)<sup>10</sup> |
| Duration of preclinical infectiousness (I<sub>p</sub> to I<sub>c</sub>; days) | 1.5 | gamma | Davies et al. (2020)<sup>10</sup> |
| Duration of clinical infectiousness (I<sub>c</sub> to R; days) | 3.5 | gamma | Davies et al. (2020)<sup>10</sup> |
| Duration of subclinical infectiousness (I<sub>s</sub> to R; days) | 5.0 | gamma | Davies et al. (2020)<sup>10</sup> |
| Incubation period (E to I<sub>c</sub>; days) | 5.5 | derived | $dE+dP$ |
| Susceptibility to infection on contact | age-dependent | derived | Davies et al. (2020)<sup>10</sup>, Davies et al. (2020)<sup>36</sup> |
| Probability of clinical symptoms on infection for age group i | age-dependent | derived | Davies et al. (2020)<sup>10</sup>, Davies et al. (2020)<sup>36</sup> |
| Relative infectiousness of subclinical cases | 50% | fixed | assumed |
| Delay from onset to hospitalisation (days) | 7.0 | gamma | Davies et al. (2020)<sup>10</sup> |
| Duration of hospitalisation in non-ICU bed (days) | 8.0 | gamma | Davies et al. (2020)<sup>10</sup> |
| Duration of hospitalisation in ICU bed (days) | 10.0 | gamma | Davies et al. (2020)<sup>10</sup> |
| Proportion of hospitalised cases that require critical care | 17% | fixed | Docherty et al. (2020)<sup>35</sup> |
| Proportion of fatal hospitalised cases that require critical care | 32% | fixed | Docherty et al. (2020)<sup>35</sup> |
| Delay from onset to death (days) | 22.0 | gamma | Davies et al. (2020)<sup>10</sup> |
| Delay from infection to death (days) | 26.0 | derived | latent period and delay from onset to death |
| Birth/deaths annually | 731,000 | fixed | ONS (2018)<sup>34</sup> |

ICU: intensive-care unit, ONS: Office for National Statistics.

In addition, we informed mortality in individuals aged <75 years per 5-year age-band using ensemble infection-fatality rates based on data on COVID-19 confirmed deaths from 45 countries,^37^ while infection-fatality rates in individuals aged ≥75 years were based on the REACT3 study from the UK^38^ (Supplementary Table 3). Our results are slightly different from earlier analyses^10^ due to using different input parameters for mortality, and the model including waning and demography. Our analysis assumed that input values can be extrapolated for the whole of the UK.

**Supplementary Table 3.** Input data of infection-fatality-rate.

| Supplementary Table 3. Input data of infection-fatality-rate. |  |  |  |
| --- | --- | --- | --- |
| Age | Median | 95% CrI (lower) | 95% CrI (upper) |
| 0-4 | 0.002% | 0.001% | 0.002% |
| 5-9 | 0.000% | 0.000% | 0.000% |
| 10-14 | 0.000% | 0.000% | 0.001% |
| 15-19 | 0.002% | 0.001% | 0.002% |
| 20-24 | 0.004% | 0.003% | 0.004% |
| 25-29 | 0.009% | 0.008% | 0.011% |
| 30-34 | 0.017% | 0.015% | 0.019% |
| 35-39 | 0.029% | 0.027% | 0.031% |
| 40-44 | 0.053% | 0.049% | 0.056% |
| 45-49 | 0.086% | 0.081% | 0.091% |
| 50-54 | 0.154% | 0.0146% | 0.162% |
| 55-59 | 0.241% | 0.229% | 0.253% |
| 60-64 | 0.359% | 0.340% | 0.379% |
| 65-69 | 0.642% | 0.610% | 0.676% |
| 70-74 | 1.076% | 1.022% | 1.132% |
| 75+ | 11.64% | 9.22% | 14.06% |

### QALY loss input data

There are currently no robust estimates of health-related quality of life associated with having (long-term) COVID-19 that used the preferred instrument in England (the EQ-5D). Consequently, the QALY losses were informed from previously published values of other respiratory infections (Supplementary Table 4), which may underestimate the health gain from preventing persistent symptoms of COVID-19 lasting several months.^26^ Once more information about COVID-19 become available these estimates could be updated later, or indeed further explored more conceptually.

QALYs lost per symptomatic case were based on ILI for 2009 H1N1 pandemic influenza in the UK,^39^ while QALY loss per non-fatal hospitalisation were based on seasonal influenza in the UK.^40^

QALYs lost per non-fatal ICU stay were based on decrements in quality of life of two studies in ICU survivors from the UK.^41,42^ The difference in utility over one year was 0.10 in the study of Griffiths et al. (2013) and ∼0.15 in the study by Cuthbertson et al. (2010), but it continued at roughly 0.10-0.15 in year 2.5 and year 5 as reported by Cuthbertson et al. (2010).^42^

QALYs lost from adverse events following immunisation (AEFI) were assumed with 1 QALD at a chance of 10%, which is roughly following another study on influenza vaccination.^43^ We did not consider longer-term or serious AEFI that may occur but are unknown yet.

QALYs lost per death were based on the most recent life expectancy in the UK as 3-year average over 2016-2018,^44^ and adjusted for age- and sex-specific QALY population norms based on the EQ-5D-3L for the UK.^45^ We also adjusted for the higher prevalence of comorbidities in individuals most likely to die from COVID-19 if infected,^46^ by reducing the QALY norms by an assumed 10% and accounting for an assumed 25% increased risk of non-COVID-19 mortality in these individuals.

To explore parameter uncertainty, we ran the epidemiological model deterministically with R0 values of 2.7 (the base case), 1.6, and 3.9.^10^ The economic model obtained 1,000 iterations using Monte Carlo sampling of the input costs and QALYs to obtain a probability distribution of outcomes. We used beta distributions for the utilities and a normal distribution for the estimated QALYs lost due to premature mortality. We explored the uncertainty using values that were ±25% of the mean value (±10% for the QALYs lost due to premature mortality) to make as few assumptions as possible on the data and variance.

**Supplementary Table 4:**
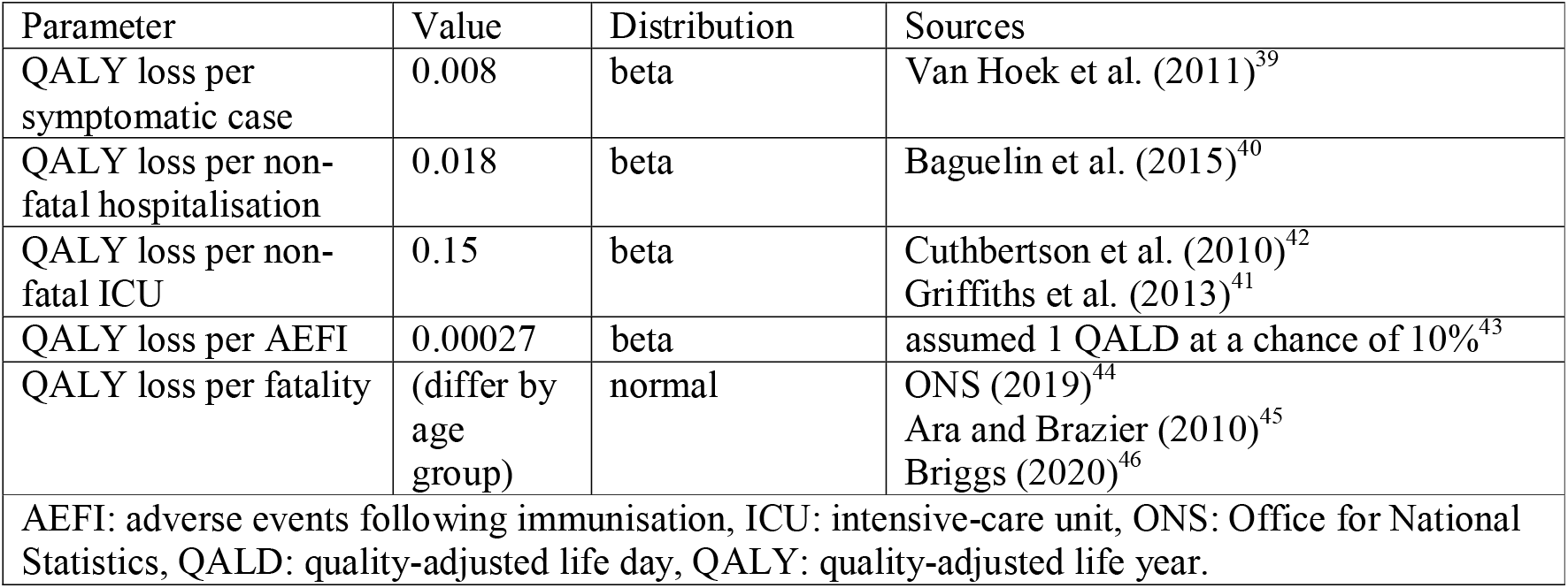
Input parameters in terms of QALYs

| Supplementary Table 4: Input parameters in terms of QALYs |  |  |  |
| --- | --- | --- | --- |
| Parameter | Value | Distribution | Sources |
| QALY loss per symptomatic case | 0.008 | beta | Van Hoek et al. (2011)<sup>39</sup> |
| QALY loss per non-fatal hospitalisation | 0.018 | beta | Baguelin et al. (2015)<sup>40</sup> |
| QALY loss per non-fatal ICU | 0.15 | beta | Cuthbertson et al. (2010)<sup>42</sup><br>Griffiths et al. (2013)<sup>41</sup> |
| QALY loss per AEFI | 0.00027 | beta | assumed 1 QALD at a chance of 10%<sup>43</sup> |
| QALY loss per fatality | (differ by age group) | normal | ONS (2019)<sup>44</sup><br>Ara and Brazier (2010)<sup>45</sup><br>Briggs (2020)<sup>46</sup> |
| AEFI: adverse events following immunisation, ICU: intensive-care unit, ONS: Office for National Statistics, QALD: quality-adjusted life day, QALY: quality-adjusted life year. |  |  |  |

### Healthcare cost input data

For the healthcare perspective, we considered the costs associated with visits to general practitioners (GPs), remote helpline calls, non-fatal hospitalisations (ICU and non-ICU), enhanced personal protective equipment (PPE), AEFI, vaccine administrations, and the vaccine costs.

Costs per GP visit were based on published unit costs in 2019,^47^ while we informed the proportion of 5% physical visits to GP practices based on published data for England from week 9-23.^48^ Similarly, remote helpline calls were approximated with calls to NHS111 in England, which were costed using the estimated costs of £12.26 per call in 2011 that we inflated to 2019 (£13.86).^47,49^ The proportion of 10% calls was again informed by the published data for England.^48^

Costs per non-ICU hospitalisation and per ICU hospitalisation were based on the NHS reference costs 2018/19.^50^ Non-ICU hospitalisations were approximated with ICD-10/HRG codes for other viral pneumonia: J12.8 (Other viral pneumonia), J12.9 (Viral pneumonia, unspecified). The base HRG code per hospitalisation for other viral pneumonia is DZ11 (PD14 for age <=18 years). ICU-hospitalisations were approximated with Adult Critical Care (activity-weighted HRG codes XC01Z-XB07Z) and Paediatric Critical Care (activity-weighted HRG codes XB01Z-XB07Z). The ICU costs are an estimate per bed-day, and we assumed a mean stay of 10 days in ICU.^10^ Of note, these hospitalisation costs are slightly lower than those published for the 2009 H1N1 influenza pandemic,^51^ which may underestimate the costs per hospitalisation and thus the cost-effectiveness of averting hospitalisations.

Costs per enhanced PPE were based on a previous study on MERS-CoV in 2015,^52^ which estimated the additional costs on enhanced PPE equipment (mask, gown, gloves, goggles) at £2.50 per patient visit. Accounting for the additional time of an estimated 15 minutes to put on and take off the PPE as well as disposal plus documentation per patient visit, at 6 visits per patient per day came at additional £29.50 for nurses and £45 for physicians, and the total costs per patient at £119 (uprated to 2019 value at £127.62).^47,52^ We conservatively estimated the costs based on the daily estimated number of inpatients, assuming one nurse caring for 8 patients.^53^ This may underestimate the true costs of PPE, which would underestimate the cost savings from avoiding hospitalisations.

For the costs per AEFI we followed the assumption made by others to use the costs of 1 GP visit per AE.^47,54^ again assuming a chance of 10%.^43^ No costs for longer-term/serious AEFI were included.

The costs of vaccine administration were based on an assumed service payment of £10.06.^55^ Although assumed to be administered via GPs, we did not account for the costs of extraordinary GP visits (i.e., we implicitly assume the vaccine to be administered as part of another visit; this assumption may be challenged given the current advise of physical distancing, and increasing the costs per vaccine was shown to be somewhat sensitive for the pessimistic vaccination scenario depending on the physical distancing scenario in sensitivity analysis).

The cost per vaccinee were conservatively assumed to be (at least) £10, which we varied in sensitivity analyses at £0-£200. In addition, the vaccination scenarios considered the public expenditures on subsidising the development of SARS-CoV-2 vaccines with £250 million by the UK government so far,^13^ which could be regarded as an extraordinary lump-sum ex-ante premium.

Additional cost factors could have been considered, including for instance an expanded testing programme or the running costs of the temporary field hospitals (estimated with approximately £15 million for the seven NHS Nightingale hospitals in England in April 2020).^56^ While cost savings may be realisable on the field hospitals, expanded testing for SARS-CoV-2 may become a fixture in the years ahead for surveillance purposes irrespective of disease activity, and may thus be regarded as a fixed sunk-cost for economic analyses.

**Supplementary Table 5:**
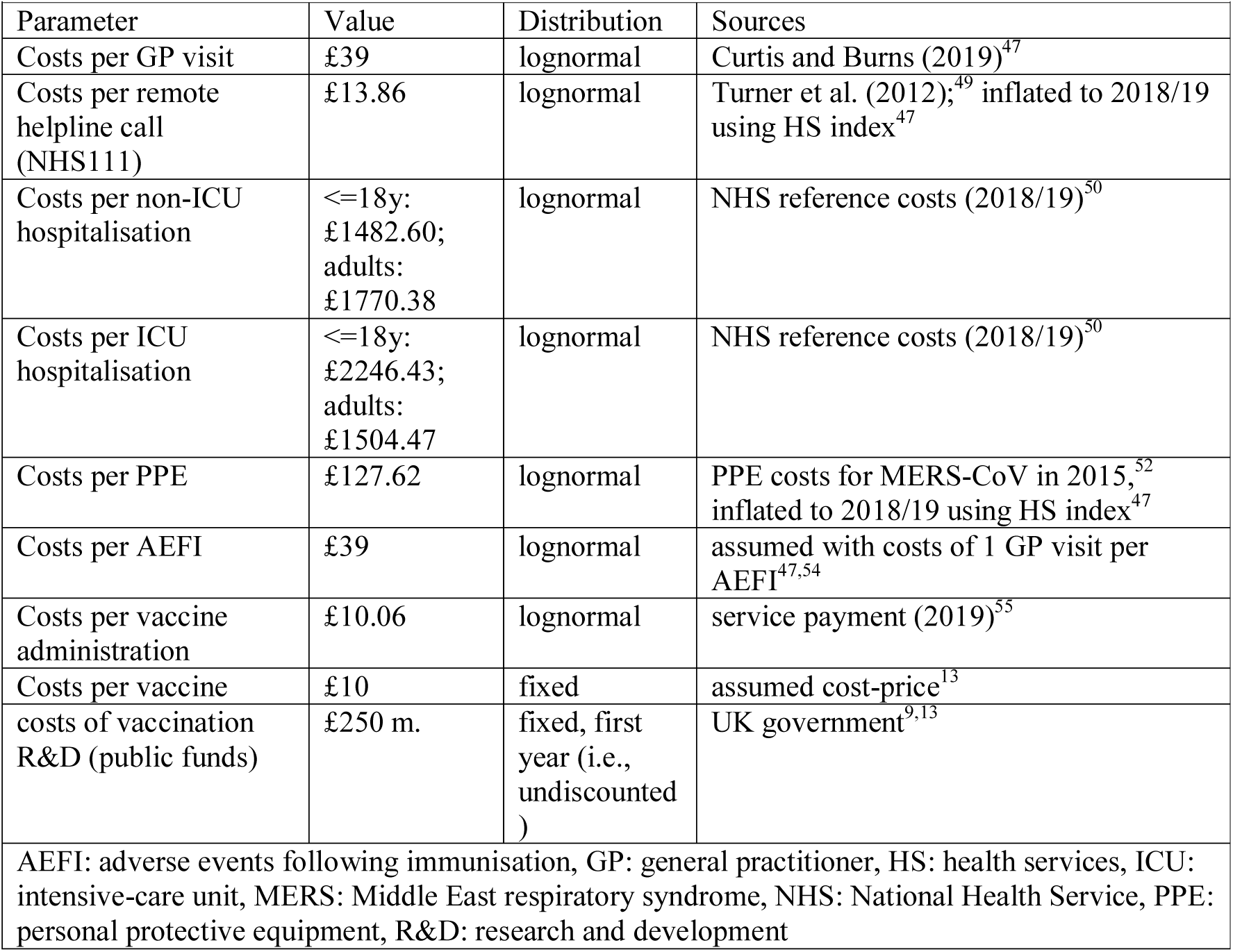
Input parameters in terms of costs

## Additional results

**Supplementary Table 6:** Results (mean, lower UI, upper UI) in terms of cases, deaths, and days spent in physical distancing per physical distancing (PD) scenario and vaccination scenario.

| Outcome | PD scenario | Vaccination scenario |  |  |
| --- | --- | --- | --- | --- |
|  |  | A | B | C |
| cases<br>(in mln.) | 1 | 147.90 (48.51, 198.73) | 113.89 (12.12, 169.80) | 47.69 (11.65, 102.98) |
|  | 2 | 141.80 (45.55, 193.42) | 108.01 (4.83, 164.64) | 44.95 (1.75, 98.08) |
|  | 3 | 21.37 (10.17, 98.25) | 14.58 (3.07, 59.76) | 11.54 (1.12, 25.57) |
|  | 4 | 30.53 (15.76, 98.85) | 22.53 (4.46, 60.87) | 32.02 (1.75, 33.06) |
|  | 5 | 37.34 (22.79, 99.14) | 25.97 (4.83, 67.96) | 32.50 (1.75, 37.71) |
|  | 6 | 43.51 (24.77, 103.45) | 32.33 (4.83, 72.94) | 32.76 (1.75, 43.90) |
|  | 7 | 48.66 (37.99, 106.02) | 37.82 (4.83, 77.34) | 33.03 (1.75, 92.21) |
|  | 8 | 53.02 (43.13, 108.66) | 43.10 (4.83, 80.59) | 33.32 (1.75, 92.02) |
| deaths<br>(in mln.) | 1 | 2.84 (0.77, 4.17) | 2.33 (0.20, 3.78) | 1.08 (0.20, 2.65) |
|  | 2 | 2.68 (0.71, 4.00) | 2.17 (0.08, 3.61) | 0.98 (0.03, 2.46) |
|  | 3 | 0.36 (0.15, 1.89) | 0.27 (0.05, 1.23) | 0.25 (0.02, 0.57) |
|  | 4 | 0.52 (0.24, 1.90) | 0.42 (0.07, 1.25) | 0.74 (0.03, 0.76) |
|  | 5 | 0.64 (0.34, 1.91) | 0.49 (0.08, 1.41) | 0.75 (0.03, 0.88) |
|  | 6 | 0.75 (0.37, 2.00) | 0.61 (0.08, 1.52) | 0.76 (0.03, 1.04) |
|  | 7 | 0.84 (0.58, 2.05) | 0.72 (0.08, 1.61) | 0.76 (0.03, 2.36) |
|  | 8 | 0.92 (0.67, 2.11) | 0.82 (0.08, 1.69) | 0.76 (0.03, 2.36) |
| days of<br>increased<br>PD | 1 | 0.00 (0.00, 0.00) | 0.00 (0.00, 0.00) | 0.00 (0.00, 0.00) |
|  | 2 | 129.00 (49.00, 149.00) | 129.00 (49.00, 149.00) | 129.00 (49.00, 149.00) |
|  | 3 | 3381.00 (892.00, 3582.00) | 2661.00 (89.00, 3571.00) | 952.00 (89.00, 2653.00) |
|  | 4 | 3183.00 (778.00, 3583.00) | 2496.00 (57.00, 3560.00) | 267.00 (49.00, 2462.00) |
|  | 5 | 3039.00 (590.00, 3576.00) | 2407.00 (49.00, 3379.00) | 261.00 (49.00, 2318.00) |
|  | 6 | 2909.00 (549.00, 3458.00) | 2267.00 (49.00, 3249.00) | 258.00 (49.00, 2134.00) |
|  | 7 | 2803.00 (245.00, 3384.00) | 2145.00 (49.00, 3135.00) | 255.00 (49.00, 278.00) |
|  | 8 | 2711.00 (111.00, 3307.00) | 2024.00 (49.00, 3052.00) | 252.00 (49.00, 280.00) |

Physical distancing scenarios: (1) no lockdown; (2) initial lockdown only; (3) PD trigger at 10/100,000 cases; (4) PD trigger at 20/100,000 cases; (5) PD trigger at 30/100,000 cases; (6) PD trigger at 40/100,000 cases; (7) PD trigger at 50/100,000 cases; (8) PD trigger at 60/100,000 cases.
Vaccination scenarios: (A): no-vaccination baseline scenario; (B): vaccination with 25% vaccine effectiveness, vaccine-induced protection of 1-year duration, and annual revaccination of 90%; (C): vaccination with 75% vaccine effectiveness, vaccine-induced protection of 10-year duration, and annual revaccination of 10% of individuals in the target population.
mln.: million, PD: physical distancing, UI: uncertainty interval.

**Supplementary Table 7:** Results (mean, lower UI, upper UI) in terms of QALYs, costs, and net monetary benefit (NMB) from the healthcare perspective per physical distancing (PD) and vaccination scenario.

| Outcome | PD scenario | Vaccination scenario |  |  |
| --- | --- | --- | --- | --- |
|  |  | A | B | C |
| QALYs<br>(in bln.) | 1 | 0.08 (0.06, 0.09) | 0.06 (0.05, 0.07) | 0.03 (0.02, 0.03) |
|  | 2 | 0.07 (0.06, 0.09) | 0.06 (0.05, 0.07) | 0.03 (0.02, 0.03) |
|  | 3 | 0.01 (0.01, 0.01) | 0.01 (0.01, 0.01) | 0.01 (0.01, 0.01) |
|  | 4 | 0.01 (0.01, 0.02) | 0.01 (0.01, 0.01) | 0.02 (0.02, 0.02) |
|  | 5 | 0.02 (0.01, 0.02) | 0.01 (0.01, 0.02) | 0.02 (0.02, 0.02) |
|  | 6 | 0.02 (0.02, 0.02) | 0.02 (0.01, 0.02) | 0.02 (0.02, 0.02) |
|  | 7 | 0.02 (0.02, 0.03) | 0.02 (0.02, 0.02) | 0.02 (0.02, 0.02) |
|  | 8 | 0.03 (0.02, 0.03) | 0.02 (0.02, 0.03) | 0.02 (0.02, 0.02) |
| costs<br>(in bln.) | 1 | 74.47 (48.69, 110.64) | 97.12 (74.11, 128.56) | 35.64 (25.91, 49.17) |
|  | 2 | 70.41 (46.04, 104.60) | 92.86 (71.18, 122.10) | 33.29 (24.36, 45.59) |
|  | 3 | 9.89 (6.48, 14.67) | 44.42 (36.86, 54.23) | 14.37 (11.54, 18.05) |
|  | 4 | 14.18 (9.29, 21.03) | 48.37 (40.14, 58.86) | 25.93 (19.44, 34.73) |
|  | 5 | 17.40 (11.40, 25.81) | 50.14 (41.55, 60.95) | 26.16 (19.59, 35.05) |
|  | 6 | 20.32 (13.31, 30.14) | 53.30 (43.81, 64.86) | 26.29 (19.68, 35.23) |
|  | 7 | 22.73 (14.89, 33.72) | 56.02 (45.77, 68.80) | 26.41 (19.76, 35.41) |
|  | 8 | 24.81 (16.25, 36.81) | 58.62 (47.88, 72.05) | 26.55 (19.85, 35.59) |
| NMB<br>(in bln.) | 1 | -402.34 (-534.56, -286.44) | -336.94 (-439.41, -242.59) | -153.84 (-199.96, -114.98) |
|  | 2 | -379.92 (-504.90, -270.32) | -313.54 (-408.79, -226.22) | -140.60 (-182.67, -104.97) |
|  | 3 | -53.12 (-70.73, -37.62) | -50.60 (-63.84, -40.55) | -37.46 (-48.17, -28.57) |
|  | 4 | -76.18 (-101.41, -53.98) | -72.01 (-91.95, -56.44) | -101.04 (-131.11, -75.92) |
|  | 5 | -93.51 (-124.47, -66.28) | -81.60 (-104.53, -63.19) | -102.27 (-132.71, -76.81) |
|  | 6 | -109.20 (-145.35, -77.42) | -98.74 (-127.03, -75.25) | -102.92 (-133.55, -77.28) |
|  | 7 | -122.19 (-162.63, -86.63) | -113.49 (-146.39, -85.63) | -103.58 (-134.42, -77.76) |
|  | 8 | -133.37 (-177.51, -94.57) | -127.60 (-164.91, -95.55) | -104.27 (-135.31, -78.25) |

Physical distancing scenarios: (1) no lockdown; (2) initial lockdown only; (3) PD trigger at 10/100,000 cases; (4) PD trigger at 20/100,000 cases; (5) PD trigger at 30/100,000 cases; (6) PD trigger at 40/100,000 cases; (7) PD trigger at 50/100,000 cases; (8) PD trigger at 60/100,000 cases.
Vaccination scenarios: (A): no-vaccination baseline scenario; (B): vaccination with 25% vaccine effectiveness, vaccine-induced protection of 1-year duration, and annual revaccination of 90%; (C): vaccination with 75% vaccine effectiveness, vaccine-induced protection of 10-year duration, and annual revaccination of 10% of individuals in the target population.
bln.: billion, NMB: net monetary benefit, PD: physical distancing, QALY: quality-adjusted life year, UI: uncertainty interval.

